# Sex- and Depression-specific Effects of Non-pathogenic CAG Repeats in *HTT*, *ATXN3* and *CACNA1A* on Sleep

**DOI:** 10.1101/2025.11.26.25341069

**Authors:** Linjun Ao, Raymond Noordam, N. Ahmad Aziz, Yuri Milaneschi, Diana van Heemst, Frits R. Rosendaal, Brenda W J H Penninx, Ko Willems van Dijk, Tamar Sofer, Heming Wang, Tariq Faquih

## Abstract

The effects of non-pathogenic cytosine-adenine-guanine (CAG) repeat sizes on sleep remain unclear, although disrupted sleep has been observed in patients with pathogenic CAG expansions in polyglutamine disease-associated genes (PDAGs), particularly in *HTT*, *ATXN3,* and *CACNA1A*. Here, we assessed the associations between CAG repeat sizes of the three genes and self-reported sleep outcomes in the Netherlands Epidemiology of Obesity study (NEO) and the Netherlands Study of Depression and Anxiety (NESDA). Sleep outcomes included excessive daytime sleepiness (EDS) and Pittsburgh Sleep Quality Index (PSQI) in NEO, insomnia score in NESDA, and sleep duration and chronotype in both. We also stratified by sex, menopausal status in women, and questionnaire-based depression score. We observed 31 associations, of which 26 were specific to women. Larger *HTT* CAG repeat sizes were associated with lower EDS risk and lower PSQI score in premenopausal women, but higher PSQI score in women with depression. CAG repeats in all three PDAGs were associated with sleep duration, with *ATXN3* showing U-shaped effects in all population groups except men. CAG repeat size in *CACNA1A* was primarily associated with chronotype in women. These findings suggest that non-pathogenic CAG repeats in PDAGs affect sleep differentially by sex and depression status.

## Introduction

Sleep is recognized as a vital biological process critical for maintaining overall health (1). Deficits in sleep duration, quality, and timing are established risk factors for numerous adverse health outcomes, including hypertension (1–3), obesity and metabolic disorders (1, 4), cardiovascular disease (1, 5, 6), and cognitive decline (1, 7, 8). Sleep health has recently been acknowledged by the American Heart Association as one of “Life’s Essential 8” factors relevant for cardiovascular disease prevention (9). Despite its importance, it is estimated that 50 to 70 million Americans chronically suffer from disturbed sleep, and the prevalence of general sleep disturbance and insufficient sleep in the Netherlands is estimated to be 32.1% and 43.2%, respectively (10). For these reasons, prioritizing research on determinants of sleep is crucial for improving health (11–13).

Genomics studies have identified genetic associations with various sleep phenotypes and revealed relevant biological pathways underlying those traits (14–17). However, tandem repeat polymorphisms, such as trinucleotide cytosine–adenine–guanine (CAG) repeats, have not been included in these efforts. This gap is due to sequencing limitations and data unavailability, despite the fact that short tandem repeats are a prominent contributor to genomic variability and comprise up to 3% of the human genome (18). CAG repeat expansions beyond the normal range are known to lead to the onset of hereditary neurodegenerative genetic diseases known as polyglutamine (PolyQ) diseases when these occur in specific genes known as polyglutamine disease-associated genes (PDAGs) (19, 20). *HTT*, *ATXN3,* and *CACNA1A* are PDAGs that are particularly interesting for sleep research. Studies in patients with Huntington disease, spinocerebellar ataxia types 3 and 6, caused by CAG repeat expansions in *HTT*, *ATXN3,* and *CACNA1A*, respectively, show symptoms of disrupted sleep, including sleep apnea (21, 22), daytime sleepiness (21, 23), and disturbances in circadian rhythm (24, 25).

Recently, CAG repeat variations within the normal range that do not lead to PolyQ disorders have been found to be associated with clinical outcomes related to sleep health. These include associations with body mass index (BMI) (26), neuropsychiatric conditions such as depression (27–29), and atherogenic lipoprotein profiles (30). In addition, as observed for BMI, CAG repeat variations within the normal range could explain a comparable amount of variation in the trait of interest as associated single nucleotide polymorphisms (SNPs) (26), and therefore are useful to explain part of the ‘missing heritability’ in GWAS.

Here, we aimed to investigate the associations of CAG repeat sizes within the normal range of PDAGs with various sleep traits. Using data from the Netherlands Epidemiology of Obesity study (NEO) and the Netherlands Study of Depression and Anxiety (NESDA), we analyzed CAG repeat sizes in the *HTT*, *ATXN3,* and *CACNA1A* genes. Sleep traits included excessive daytime sleepiness (EDS) and Pittsburgh Sleep Quality Index (PSQI) from NEO, Women’s Health Initiative Insomnia Rating Scale (WHIIRS) from NESDA, and sleep duration and chronotype (morningness and eveningness) from both cohort studies.

## Results

### Sample characteristics

In the NEO study, we included 3961 individuals (51.1% women) with genotyping and sleep questionnaire data. These individuals had a median age of 56 years (interquartile range [IQR]: 51-61 years) and a mean BMI of 29.7 kg/m^2^ (standard deviation [SD]: 4.85 kg/m^2^). Among the included individuals, based on the Inventory of Depressive Symptomatology Self-Report (IDS-SR) questionnaire, 1084 (27.4%) had an IDS score ≥ 14. In NESDA Visit 1, for the outcome of WHIIRS, a total of 2165 individuals (66.8% women) were included, with a median age of 44 years (IQR: 31-54), a mean BMI of 25.5 kg/m^2^ (SD: 4.8), and IDS score ≥ 14 in 62.1%. In NESDA Visit 3, for the outcome of sleep duration and chronotype, a total of 1788 individuals (66.6% women) were included, with a median age of 45 years (IQR: 32-55 years), a mean BMI of 25.6 kg/m^2^ (SD: 4.8), and IDS score ≥ 14 in 44.6%. The distribution of CAG repeat sizes for each allele in each gene was similar across cohorts and sex groups. Detailed population characteristics for the included studies are provided in Table 1.

**Table 1.** Characteristics of included study population in NEO and NESDA.

### CAG repeats associations with self-reported sleep traits

A study summary is shown in Figure 1. Briefly, associations between CAG repeat sizes in each gene and sleep traits were assessed using regression models in the whole population and sex-stratified groups, with further stratification by menopausal status in women. Additional sensitivity analyses, such as interaction and stratification by IDS score, were also performed. Models incorporated interaction or quadratic terms of alleles when non-linear effects were present. Analyses were performed separately in each cohort, with results for sleep duration and chronotype meta-analyzed.

**Figure 1.**
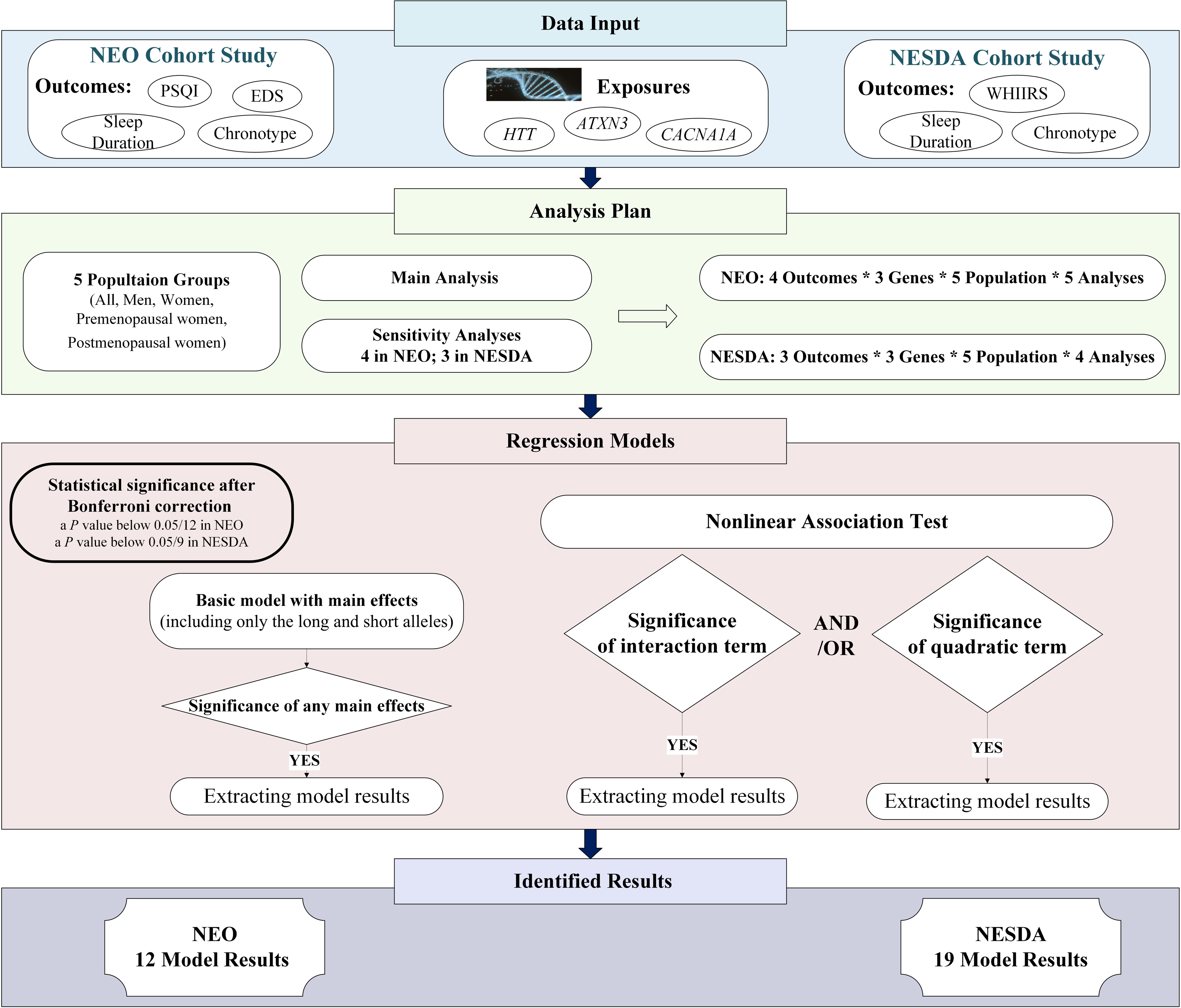
A flowchart for the study design and results. Abbreviations: NEO, Netherlands Epidemiology of Obesity; NESDA, Netherlands Study of Depression and Anxiety; EDS, excessive daytime sleepiness; PSQI, Pittsburgh Sleep Quality Index; WHIIRS, Women’s Health Initiative Insomnia Rating Scale

Overall, we observed 31 statistically significant associations (*P*-value < 0.0042 in NEO and *P*-value < 0.0056 in NESDA; Bonferroni adjusted), including 12 in NEO and 19 in NESDA across the main and sensitivity analyses. A simple summary of the results is shown in Figure 2. We observed associations for sleep duration and chronotype in both studies, as well as EDS and PSQI in NEO. No associations were found for WHIIRS scores in any models in NESDA. However, potential associations with nominal significance (*P*-values < 0.05) were observed between the longer *HTT* CAG repeats and higher WHIIRS scores in men and lower WHIIRS scores in women (Supplementary Data 1). Full results of the 31 associations are provided in Table S1, of which the model coefficients from the main analyses are shown in Figure 3. Most of the observed associations were specific for women (7 associations) and women subgroups (13 in premenopausal women and 6 in postmenopausal women). Four associations were observed in the NESDA overall population, and one in men (NEO). Consistent with those findings stratified analyses, interactions of PDAGs with depression, sex and menopause status in women (Table S2) were observed.

**Figure 2.**
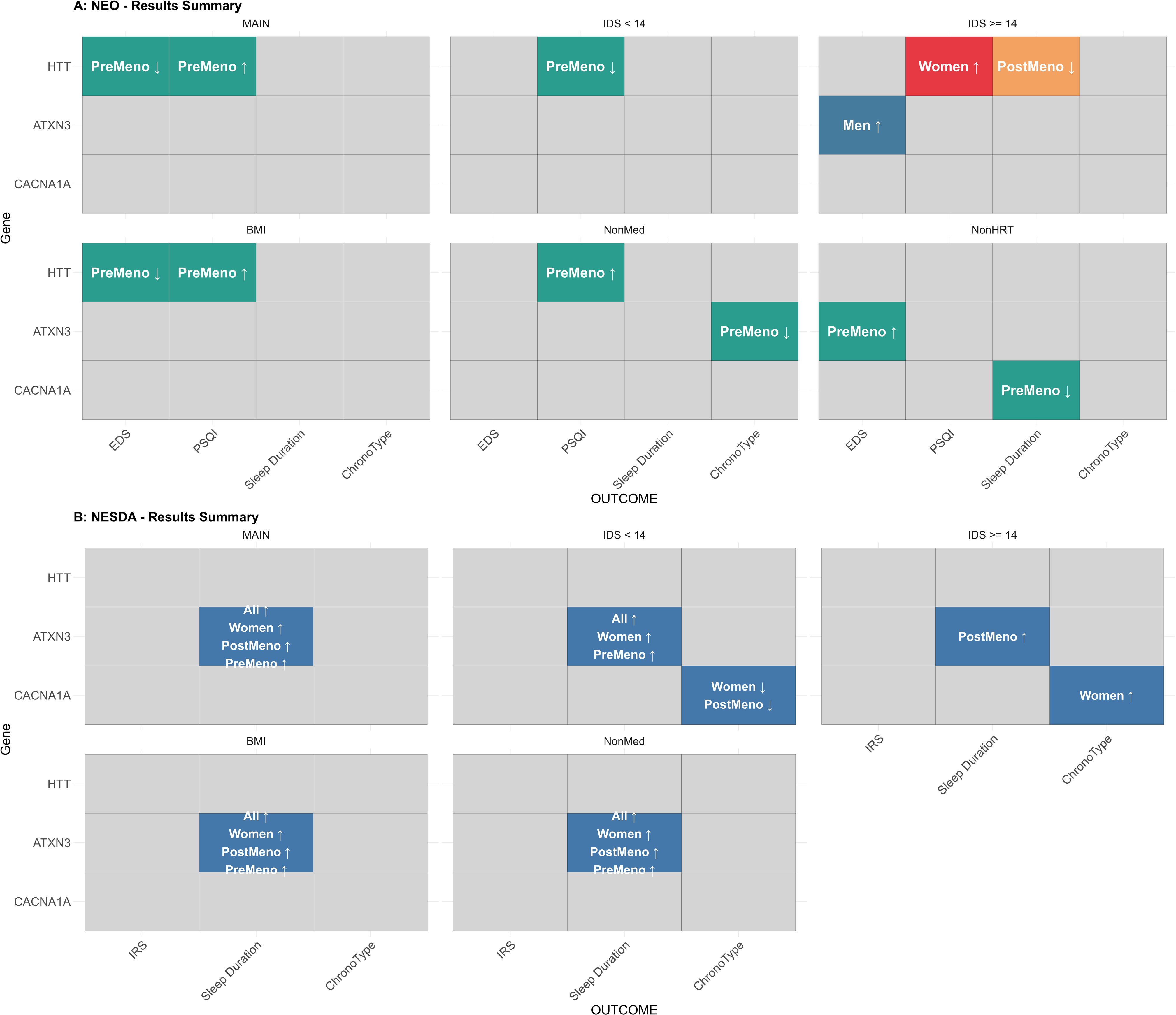
Overview of the associations identified in the main and sensitivity analyses with the effect directions of long alleles. Each grid indicates whether a significant association was identified for the corresponding sleep outcome and PDAG after Bonferroni correction. Non-grey colors represent significant associations observed in at least one population group, while grey indicates no significant associations across all groups. Grids under the label ‘MAIN’ shows results from the main analyses, while the grids labeled ‘IDS <14’, ‘IDS >= 14’, ‘BMI’, ‘NonMed’, ‘NonHRT’ shows results from the corresponding sensitivity analyses. The x-axis indicates the outcomes from each cohort study, the y-axis indicates the studied PDAGs, and the arrows indicate the effect direction of the long allele in each identified model. In the subplot of A, different colors indicates different groups in which the associations were identified.

**Figure 3.**
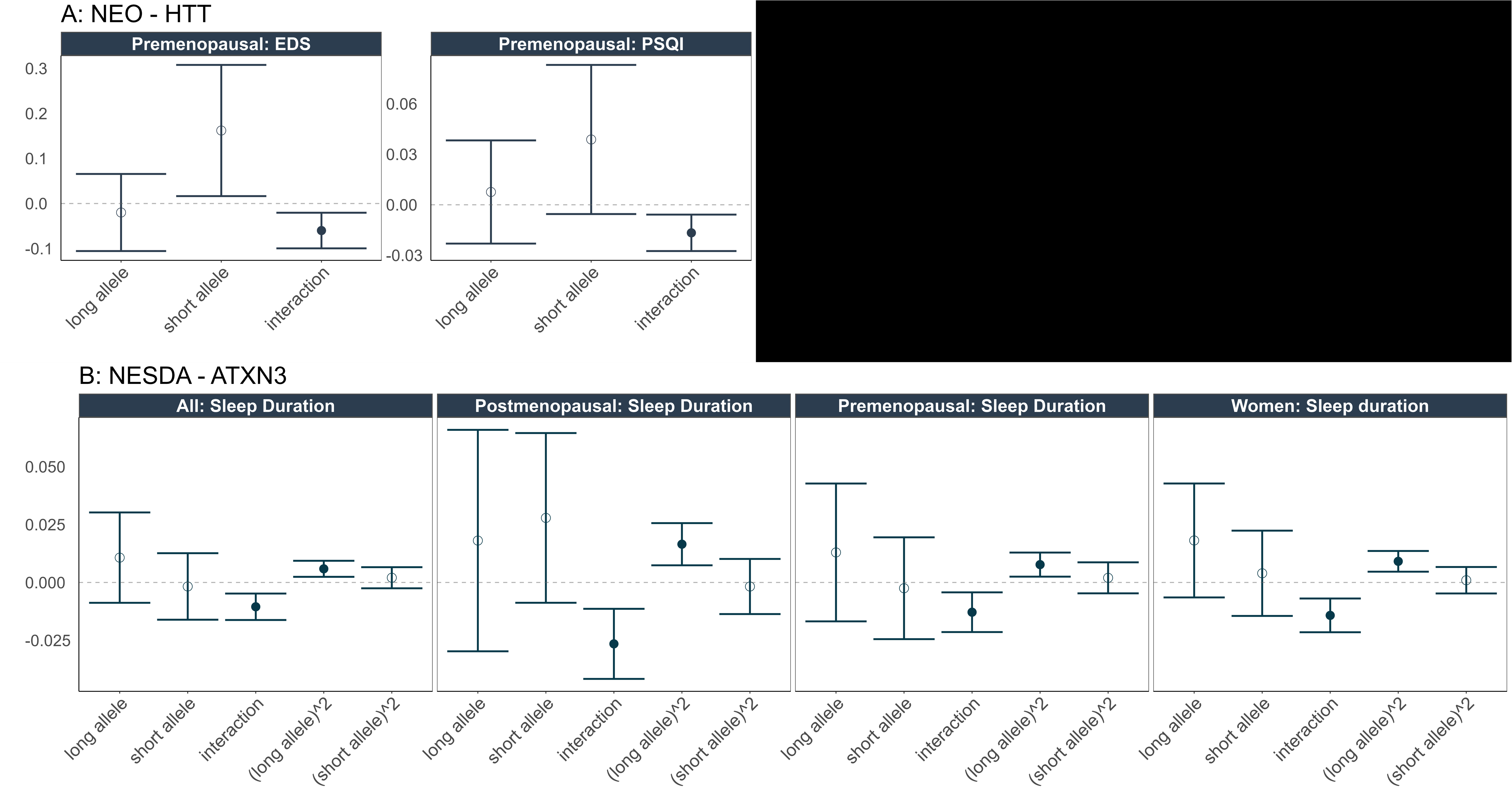
Identified associations from the main analyses. The x-axis represents the CAG repeat allele variables included in each model, and the y-axis shows the estimates in β coefficients with 95% confidence intervals. Solid points indicate statistically significant effects after Bonferroni correction, whereas empty points indicate non-significant effects.

#### Excessive daytime sleepiness

In the NEO study, in premenopausal women, we observed an association between the interaction of the *HTT* long allele and *HTT* short allele with lower odds of EDS, with an explained variance of 1.9% by the gene. This association remained consistent after adjusting for BMI. We further observed a nonlinear association between longer CAG repeats in the *ATXN3* gene (with an interaction between the two alleles) and higher odds of EDS in men with IDS ≥ 14. This association had an explained variance of 7.8%. In the sensitivity analysis of premenopausal women, excluding those with a history of hormonal replacement therapy (HRT), a linear association (no interaction effects) between the longer *ATXN3* CAG repeats and higher odds of EDS was observed, with an explained variance of 2.4% by the *ATXN3* gene. This linear positive effect (*P*-value = 0.005) remained in the general premenopausal women but was not statistically significant after Bonferroni correction (Supplementary Data 1).

#### Pittsburgh score (PSQI)

Analyses of the PSQI score showed several associations with the *HTT* CAG repeats in women. In the main analyses, for premenopausal women, longer CAG repeats in *HTT* were associated with higher PSQI scores, with an interaction between the two alleles, explaining 1.7% of the variance. The sensitivity analyses for premenopausal women, including BMI-adjustment and restriction to individuals not taking sleep medications, showed similar results. In addition, an association between the *HTT* long allele and higher PSQI was observed for women with an IDS score ≥ 14, regardless of menopausal status, explaining 1.9% of the variance. In contrast, when the IDS score < 14, only the interaction between *HTT* alleles was associated with lower PSQI scores in premenopausal women, explaining 4.4% of the variance.

#### Sleep Duration

Figure 4 shows the associations between *ATXN3* and sleep duration from the main analyses in each cohort and the corresponding meta-analyses. Across four groups (all participants, women, premenopausal women, and postmenopausal women), we consistently observed longer sleep duration associated with the quadratic term of the *ATXN3* long allele, and shorter sleep duration associated with the interaction of short and long alleles in the *ATXN3* gene (except in the post-menopausal women). Additional results in Figures S3 showed an association between the interaction of the *HTT* long and short alleles and shorter sleep duration in postmenopausal women. Sensitivity analyses excluding individuals with HRT showed a nonlinear association between *CACNA1A* and sleep duration in NEO premenopausal women (Table S1). Of these observed associations, the explained proportion of variance in sleep duration was 2.4% for *HTT* and 3.4% for *CACNA1A*, while *ATXN3* explained between 1.0% and 5.6% in the main analyses; complete explained variance (R^2^) results are presented in Table S1.

**Figure 4:**
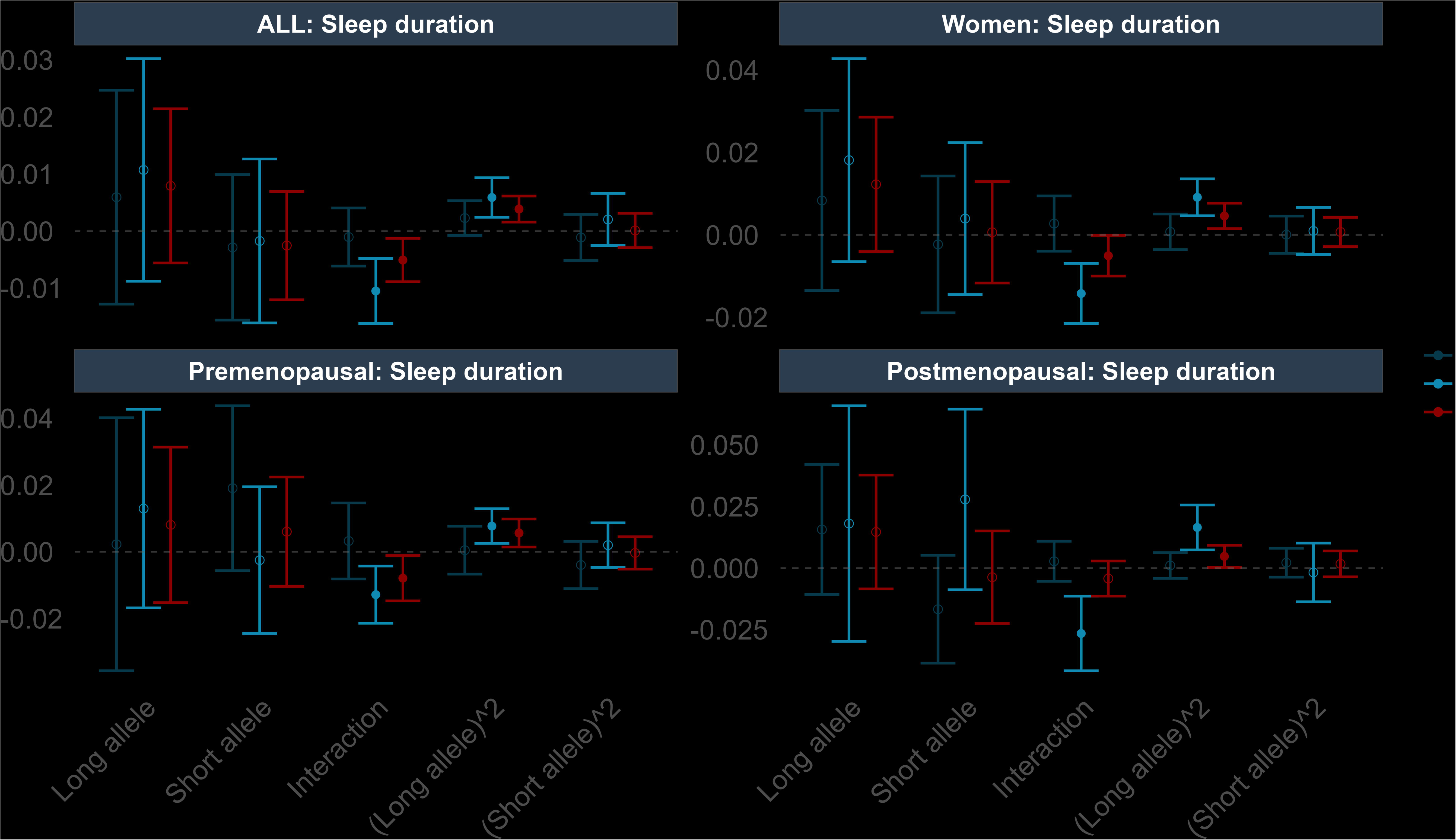
Meta-analyses for the main results of *ATXN3* with sleep duration. The x-axis represents the CAG repeat allele variables included in each model, and the y-axis shows the estimated effect sizes (β coefficients) with 95% confidence intervals. Solid points indicate statistically significant effects after Bonferroni correction, whereas empty points indicate non-significant effects. Results from the NEO study, NESDA study and meta-analyses are shown in blue, green and red, respectively.

#### Chronotype

Analyses results for chronotype in each cohort and the corresponding meta-analyses are presented in Figures S4, showing main associations with *CACNA1A*. In individuals with an IDS score ≥ 14, a positive association between allele interactions in *CACNA1A* and morning chronotype was observed in women. In individuals with an IDS score <14, the short allele in *CACNA1A* was associated with an increased odds of morning chronotype and evening chronotype in women and postmenopausal women, respectively. In addition, the longer CAG repeats in the *ATXN3* gene were associated with reduced odds of morning chronotype in premenopausal women (restricted to women not taking sleep medications). Among these observed associations, the proportion of variance in chronotype explained by *CACNA1A* ranged from 0.91% to 3.4%, while *ATXN3* accounted for 1.9% (Table S1).

### Estimated effects of CAG repeat length

The overall effects of CAG repeat sizes in the long allele on sleep outcomes are presented in Figure 5 (from main analyses) and Figure S5-6 (from sensitivity analyses). Based on the observed associations from the main analyses in NEO, larger CAG repeat sizes in the *HTT* long allele were predicted to reduce the log-odds of EDS and PSQI score in premenopausal women (Figure 5). In NESDA, the overall effect of the *ATXN3* long allele, including its interaction effect with the short allele and the effect from its quadratic term on sleep duration, showed a U-shaped pattern across different population groups except in men (Figure 5). A higher total effect was observed at higher absolute deviations from the mean, specifically, at both ends of the long allele CAG repeat range. Notably, in NEO, women with IDS ≥ 14 showed higher PSQI scores with longer *HTT* CAG repeats (Figure S5), which was opposite to the main findings. In NESDA, for individuals with IDS ≥ 14, the U-shaped pattern remained only in postmenopausal women with stronger effects (Figure S6).

**Figure 5.**
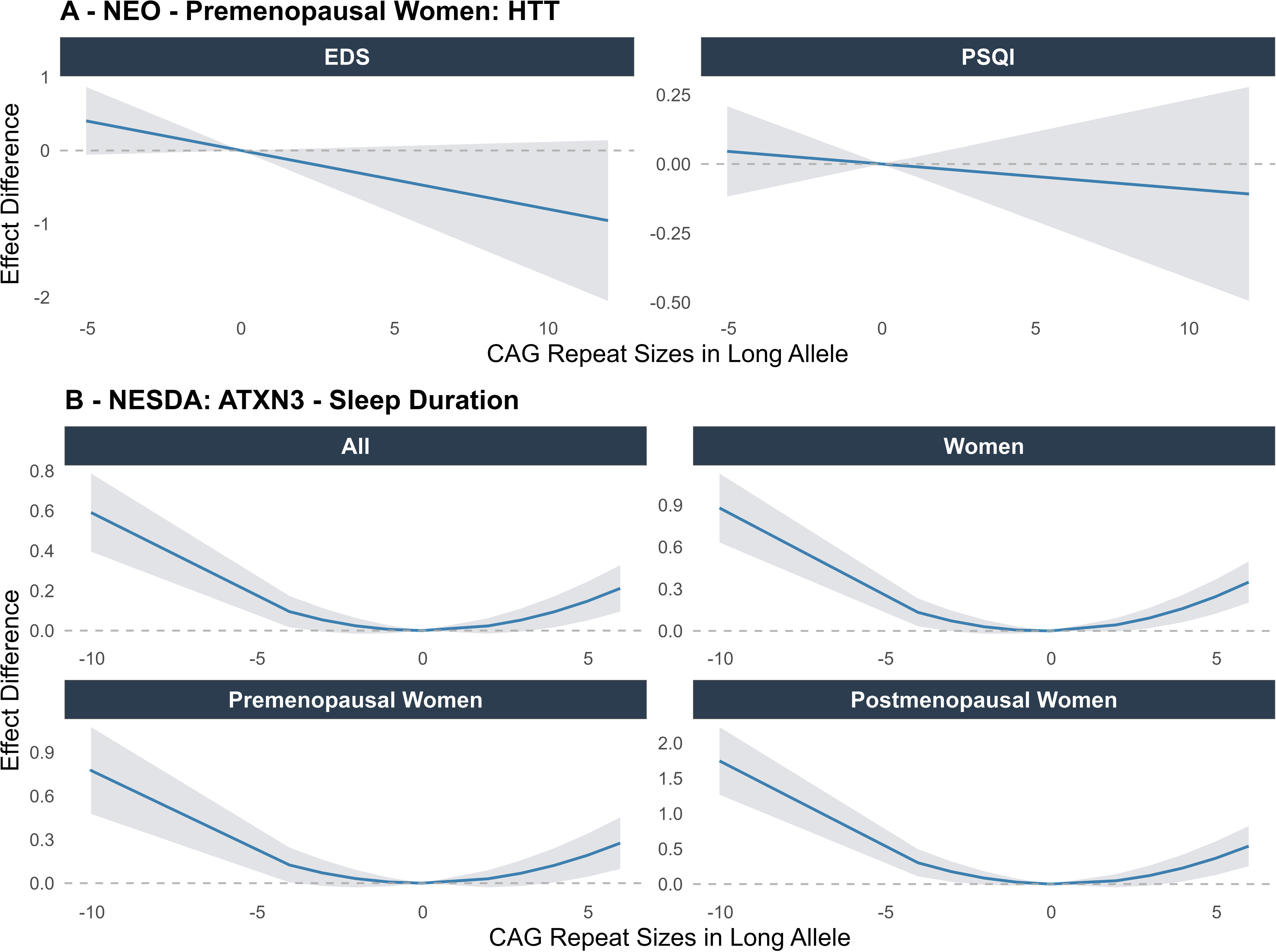
Estimated overall effects of CAG repeat sizes in long allele on the sleep outcome from main analyses. Subplot A shows the estimated overall effects of *HTT* long allele on EDS and Pittsburgh score (PSQI) for premenopausal women. Subplot B shows the estimated overall effects of *ATXN3* long allele on sleep duration for different population groups. The x-axis represents the centered CAG repeat values of the long allele, and the y-axis shows the estimated effects (lines) with 95% confidence intervals (shaded area). The outcomes of sleep duration and PSQI are scaled, with predicted effects expressed in standard deviations, whereas the predicted effects for EDS are presented as log odds ratios.

### Mediation analysis

First, normal-range CAG repeat sizes showed linear associations with the baseline dichotomized IDS scores, with an effect size of 0.14 (SE = 0.06, *P*-value = 0.01) for the *HTT* short allele in NEO premenopausal women, and 0.05 (SE = 0.02, *P*-value = 0.008) for the *ATXN3* longer allele in all NESDA participants (Table 2). Full mediation results are presented in Table S3, with no significant mediation effects of IDS observed.

**Table 2.** Significant associations between PDAG CAG repeat sizes and the dichotomized Inventory for Depressive Symptomatology score.

## Discussion

### Overall results

In this study, we examined the associations between non-pathogenic CAG repeat sizes in three PDAGs (*HTT, ATXN3,* and *CACNA1A)* with various sleep traits in two Dutch cohorts. We found that larger CAG repeat sizes in the *HTT* long allele reduced the odds of EDS but increased PSQI score in premenopausal women. However, when taking into account the interaction effects between the long and short alleles, the estimated effects of 1-CAG repeat over the mean in both alleles of *HTT* lowered the odds of EDS and the PSQI score. All three PDAGs were associated with sleep duration, with the *ATXN3* long allele showing a nonlinear association with increased sleep duration across all population groups, except in men.

The sensitivity analyses for depression status provided further insights as well. For example, larger CAG repeat sizes in *HTT* long allele increased the PSQI score in depressed but not in non-depressed women, consistent with the observed interaction between IDS score and *HTT* in women. Similarly, larger CAG repeat sizes in *ATXN3* increased EDS in depressed but not in non-derepressed men, consistent with the observed interaction between IDS score and *ATXN3* in men. Moreover, our study did not observe mediation effects of depression, suggesting that depression is likely to act as an effect modifier on the CAG-sleep associations.

Furthermore, the present study provided evidence that the normal-range CAG repeat variation in the PDAGs could account for a large portion of the genetic explained variance for sleep traits, particularly compared to SNPs. For instance normal-range CAG repeat size variation in *ATXN3* explained 1.9% of the variance in EDS for premenopausal women and 2.4% of the variance in EDS for men with IDS ≥ 14 (Table S1); whereas the cumulative explained variance of all 42 identified SNPs for EDS (based on a UKB GWAS) only explained 0.1% of the variance (16).

### *HTT* and Sleep

Previous studies have reported that the overexpansion of CAG repeats and mHTT protein is associated with sleep and circadian rhythm disruption in patients with Huntington disease (31), Drosophila models (32, 33), and R6/1 and R6/2 mice models (31, 34). These effects may result from disrupted circadian rhythmicity, reduced neural activity in the suprachiasmatic nucleus (34, 35), delayed melatonin secretion onset (36), and altered expression of clock genes such as *Period2* (*PER2*) in mice, *period* (*per*) (32, 37), *timeless* (*tim*) (32), and clock (*clk*) (32) in Drosophila. In addition, in HD patients, insomnia (31, 34, 38), delayed sleep phase (31), and fragmented sleep (31, 34, 38) have been reported. Consistent with these previous findings, our study showed that the long allele in *HTT* was associated with decreased sleep quality, reflected by higher PSQI scores among premenopausal women and depressed women. However, the *HTT* long allele was associated with lower odds of EDS. This is in line with some studies reporting that daytime sleepiness may not be significantly affected in HD patients (12.7% in HD cases compared to 7.9% controls) (24). In addition, based on our study, it is likely that the effects of *HTT* on EDS and PSQI are varied by the interaction between the long and short alleles until the CAG repeat size exceeds the pathogenic range. Moreover, although no associations between *HTT* and insomnia were found after multiple testing correction in our study, a nominal association between longer CAG alleles and WHIIRS scores in men was observed. Future studies are warranted to provide additional evidence based on a larger sample size and detailed insomnia data.

### *ATXN3* and Sleep

Spinocerebellar ataxia type 3 (SCA3), also known as Machado-Joseph disease, is one of the neurodegenerative diseases caused by an abnormal expansion of the polyQ tract of the *ATXN3* gene (39). In a study with 38 cases of SCA3 and 40 controls, the frequency of REM sleep behavior disorder and restless leg syndrome was significantly higher in the SCA3 group, but there was no difference between the groups with regard to EDS (40). Another study observed a similar finding as well (41). A previous study (42) also reported that mutant *ATXN3* can differentially affect the transcription of core clock genes like *Bmal1* and *Per2* in mice, and put forth *ATXN3* as a potential drug target for SCA3 and circadian disruption. Consistent with these previous studies, we observed evidence for a role of *ATXN3* in sleep duration and chronotype, which was mainly present in women. In contrast, we also observed evidence for a role of *ATXN3* in EDS in depressed men and premenopausal women. Our study thus indicates that the effects of *ATXN3* on sleep disorders vary by sex and menopausal status.

### *CACNA1A* and Sleep

Furthermore, our study mainly observed associations between *CACNA1A,* a gene encoding the alpha1A subunit of the Cav2.1 calcium channel (43, 44), and chronotype in women across both IDS <14 and IDS ≥14 groups, but a significant interaction effect between long and short alleles was found only in the IDS ≥ 14 group. This evidence was partly consistent with previous research, which showed significant interactions between *CACNA1A* and Psychosocial Risk Score for narcolepsy (45). In addition, previous studies showed the associations between genetic variants in *CACNA1A* and severe neurological impairment with sleep apnea symptoms (46) and EDS (16). In line with this evidence, our study observed a potential nonlinear association of *CACNA1A* with EDS in women, which was not presented in our results due to the strict criteria for statistical significance. Interestingly, our study also showed that the effects of *CACNA1A* on chronotype could differ by menopausal status, with increased odds of morning chronotype in non-depressed women, but of evening chronotype in non-depressed postmenopausal women. Overall, the combined evidence may suggest that *CACNA1A* could play a role primarily in women’s sleep health and interacts with depression severity and menopausal status.

### Dopamine, Depression, and Sex Differences

Our findings suggest that the association between longer CAG repeats and sleep outcomes may be modulated by menopausal status. A potential hypothesis is that this association is driven by a hormone-dopamine interaction. Disturbed dopamine signaling is a consistent pathological feature of both HD (47) and SCA3 (48). Dopamine also plays an important role in various sleep aspects (49–51), such as slow-wave sleep and REM sleep (50, 52). In addition, estradiol, a form of estrogen, has been shown to increase dopamine synthesis and release (53), suggesting that hormonal fluctuations can modulate dopaminergic signaling. Menopause, characterized by a decline in estrogen levels, may therefore contribute to reduced dopaminergic activity, and further could disrupt sleep architecture and lead to sleep disorders. Therefore, the observed effects varied by menopausal status in our study suggest a potential vulnerability period when hormonal shifts might amplify the impact of CAG repeat expansions. Furthermore, both our findings and prior research (54) showed the association between CAG repeat size and depression. It is well established that dopamine is closely linked to mood regulation and that lower dopamine levels and dopaminergic neurotransmission are associated with depression as well (55, 56). The higher prevalence of depression in perimenopausal and postmenopausal women (57) may thus be partly due to menopause-related declines in dopamine.

Taken together, these findings point to a potentially complex, dynamic interaction between CAG repeat sizes in PDAGs, sex hormones, dopamine signaling, and sleep regulation. Therefore, the observed sex-, menopause-, and depression-specific effects in the present study may be partly explained by the roles of dopamine in regulating sleep and depression, and its interactions with sex hormones.

### Strengths and limitations

A major strength of the present study is that it is the first to investigate the effects of normal-range CAG repeat variation in PDAGs on sleep health using a comprehensive analytical framework in the general population. Second, the genotyping methodology used in the two cohorts ensured high specificity and sensitivity by using specifically designed primers to sequence the CAG repeats regions in each PDAG. Third, although sleep disturbances have been reported in HD patients, the effects of CAG repeats on sleep remain uncertain and underexplored. Our study is the largest study thus far to examine these repeats in relation to sleep health in a large population, providing new genomic insights into sleep regulation.

However, some limitations should also be taken into account when interpreting our study. First, although we investigated various sleep traits and disorders, limited overlap in outcomes was available in both cohorts. Other common sleep conditions, such as sleep apnea, and more objectively determined sleep data, such as those from actigraphy or polysomnography measures of sleep, were also not available in this study. Second, the smaller sample size in subgroups of the population could lead to inflated R² values due to overfitting. Third, the full interpretation of the effects was not straightforward due to substantial interactions between the alleles. Nevertheless, we used the estimation models to improve interpretations. Lastly, this study included only participants of European ancestry, which limits the generalizability of the findings to other populations.

In conclusion, the present study identified novel associations of normal-range CAG repeat sizes in *HTT*, *ATXN3*, and *CACNA1A* with various sleep traits, which were differential by sex groups and depression status. Our findings also suggest that a considerable amount of variation in sleep traits could be explained by specific CAG repeats. In the future, larger studies examining these CAG repeats in conjunction with SNPs in a diverse population would provide added value in exploring sleep health and its pathophysiology.

## Methods

### Study Designs

#### The Netherlands Epidemiology of Obesity study

The NEO study is an ongoing, population-based prospective cohort study investigating men and women aged 45-65 years living in the greater Leiden area of the Netherlands. Recruitment took place from September 2008 to September 2012 at the NEO study center located at Leiden University Medical Center, enrolling a total of 6671 participants with an oversampling of participants self-reporting a BMI of 27 kg/m^2^ or higher. The study was approved by the medical ethical committee of the Leiden University Medical Center (58). In the present study, the main inclusion criteria were participants of European ancestry with complete CAG data and self-reported sleep-related outcome data, and participants with a frequency for any CAG repeat number greater than 10 to reduce the influence of outliers. Details about the inclusion criteria are presented in Figure S1.

#### Netherlands Study of Depression and Anxiety

The NESDA study is an ongoing longitudinal cohort focusing on the long-term course and consequences of depressive and anxiety disorders. NESDA was approved by the Medical Ethical Committee of the VUmc (reference number 2003/183), and all participants provided written informed consent (59). Baseline recruitment of 2981 participants, which consisted of 1701 persons with a current (six-month recency) diagnosis of depressive or anxiety disorders, took place between 2004 and 2007 from the general population, general practices, and secondary mental health centers (59). The inclusion criteria in NESDA were the same as those in NEO, as detailed in Figure S2.

### Genotyping

Due to the technical limitation of next-generation short-read sequencing to accurately call deoxyribonucleic acid repeat sequences (60), a multiplex polymerase chain reaction method was developed using a TProfessional thermocycler (Biometra, Westburg) with labelled primers to genotype the CAG repeat sizes in the *HTT*, *ATXN3*, and *CACNA1A* alleles. Further details regarding the genotyping and methodology have been described in detail previously (54).

### Sleep Outcomes

In NEO, participants provided self-reported information for the Epworth sleepiness scale (ESS) questionnaire (61), sleep quality (Pittsburgh Sleep Quality Index (PSQI) (62)), and sleep duration at the baseline survey. The ESS questionnaire (61) is used to assess the tendency to doze off during the day, with a score of 10 and higher indicating excessive daytime sleepiness (EDS). The PSQI (62) is a score to assess the overall quality of sleep and ranges from 0 to 21, with higher scores indicating greater sleep problems, and scores above 5 generally suggesting significant sleep difficulties. The outcome of self-reported sleep duration is in hours. Individuals with a sleep duration of less than 4 hours were excluded. Chronotype (63) was derived based on the self-reported typical bedtime and waking times, from which the midpoint of sleep (MPS) was calculated. Participants with an MPS less than 0.5 (very early type) or greater than 8 (very late type) were excluded to avoid statistical bias due to extreme values. Based on the quintile of MPS, participants were then divided into three groups: morning chronotype (the first quintile), evening chronotype (the last quintile), and intermediate chronotype (the remaining 60% of participants). Overall, we analyzed the outcomes of EDS, the total PSQI score, sleep duration, and chronotype.

In NESDA, the sleep traits were collected from two different visit times (Visit 1 and Visit 3). In brief, participants provided self-reported questionnaires regarding insomnia symptoms at Visit 1, and completed questionnaires regarding their sleep duration (64) and the Munich Chronotype Questionnaire (65) at Visit 3. The score of insomnia symptoms was based on the Women’s Health Initiative Insomnia Rating Scale (WHIIRS), which includes 5 different aspects of sleep in the past four weeks on a 5-point scale ranging from 0 to 20 (66). Both the total score and a dichotomous variable with 9 as a cut-off value for insomnia symptoms were analyzed in the present study. The sleep duration outcome was computed as the average weekly sleep duration, specifically (5*sleep duration work days + 2 * sleep duration on free days)/7 (64). The definition of chronotype is similar to the definition in NEO, both of which were based on the MPS. However, the calculation of MPS from NESDA was further improved to correct for the ‘sleep debt’ accumulated during the work days (65). Based on the quintile of the improved MPS, participants were defined as the morning chronotype (the first quintile), evening chronotype (the last quintile), and intermediate chronotype (the remaining 60% of participants).

### Other variables

Other variables were also collected from both cohort studies, including basic characteristics (age and sex), self-reported menopausal status (pre- and post-menopausal status), BMI, use of sleep medications, and self-reported depression status. The depression status was assessed using the Inventory of Depressive Symptomatology Self-Report (IDS-SR) questionnaire (67). Since the sleep outcomes in NESDA were from two different visit times, the other variables mentioned here were repeatedly collected at the two visit times in order to perform analyses for the corresponding sleep outcomes. Further details on the variables included in the study are provided in the supplementary material.

### Statistical Analysis

#### Main analyses

For each gene (*HTT*, *ATXN3,* and *CANCA1*), we identified the relatively long and short alleles in each cohort. This was done as the two alleles can have independent effects. The number of repeats in each allele was then centered on their respective mean within each cohort. In each cohort, we then performed regression models for the CAG repeat sizes in each gene, with linear regression for the continuous outcomes, quasi-binomial logistic regression for the dichotomized outcomes, and multinomial logistic regression for the outcome of chronotype with three outcome values.

Firstly, a basic model was performed including only the (centered) lengths of the long and short alleles for each gene and outcome. Secondly, to account for possible nonlinear associations (26, 54), we tested whether an interaction between the long and short alleles and/or the quadratic term of each allele should be included. If evidence for multiplicative interaction or quadratic effects was found, the corresponding term was added to the model. Note that evidence for a quadratic effect had to meet three criteria after correction for multiple testing: (1) the quadratic term should be statistically significant; (2) at least one of the main or interaction effect should be significant; and (3) inclusion of the quadratic term should improve model fit based on Akaike Information Criterion (AIC) (68).

All analyses in the NEO study were weighted to account for the oversampling of overweight individuals. In each cohort study, regression models were performed in all participants and sex-stratified groups (women and men), with further stratification by menopausal status in women (premenopausal and postmenopausal women). In addition, considering the multiple testing, Bonferroni correction was applied based on the number of exposures and outcomes, not subgroup analyses, to avoid an overly conservative threshold. The statistical significance was therefore defined as a *P*-value < 0.0042 (0.05/12, where 12 represents 3 PDAGs * 4 outcomes) in NEO and a *P*-value < 0.0056 (0.05/9, where 9 represents 3 PDAGs * 3 outcomes) in NESDA. Furthermore, we calculated the incremental coefficients of determination (R^2^), defined as the difference between the R^2^ of the conducted regression model and that of the corresponding model without any alleles, to assess the variance explained by the corresponding PDAGs. Specifically, we used adjusted-R^2^ for linear regression models and pseudo-R^2^ for (multinomial) logistic regression models to obtain analogous measures of model fit in the context of likelihood-based estimation. The main study design is presented in Figure 1. All analyses were performed in R (version 4.3.1), where the ‘survey’ R package was used for weighting analyses in NEO.

#### Sensitivity analyses

Sensitivity analyses included stratification by IDS score with a cutoff of 14 (≥14 indicating depression status), adjustment for BMI, exclusion of participants using sleep medication, and exclusion of women using hormonal replacement therapy (HRT) in NEO only due to data availability. In addition, given well-known differences in sleep health by sex and depression (69), and well-known associations between PDAGs and depression (54), we first performed these stratified analyses described above and subsequently complemented them with formal interaction analyses here. Specifically, in each cohort, we tested interactions of CAG repeat alleles with sex and IDS in all participants, with IDS in women and men separately, and with menopausal status in women only.

#### Meta-analysis

Meta-analyses were performed for the results of self-reported sleep duration and chronotype, the two outcomes available in both studies. For each gene and each outcome, the overall estimates of all alleles and higher-order terms from the two studies were estimated using fixed-effects multivariate meta-analysis, implemented with ‘metafor’ package in R (version 4.3.1).

#### Estimation of the effect of CAG repeat length

We estimated the effect of a range of CAG repeat sizes in the long allele on sleep outcomes, while fixing the length of the short allele to 1, in the two populations. We added the effect of the interaction term and the quadratic term (if applicable). Accordingly, the estimated effect of the long allele for each potential length value *Length_L_* is given by:

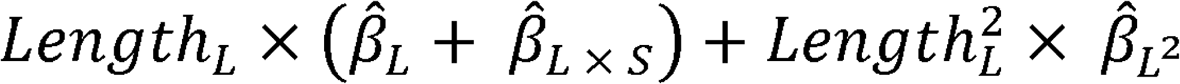

Where 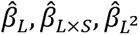 are the estimated effects of the long allele, of its interaction with the short allele length (if applicable), and of the quadratic terms of the long allele (if applicable), respectively. In addition, the delta method was used to approximate the variance of the estimated effect for each potential length value *Length_L_*, which is detailly described in supplementary material.

#### Mediation analysis

Based on the sensitivity analyses for IDS score, we further investigated whether IDS score mediated the associations between CAG repeat sizes and sleep traits. Mediation analyses focused on the associations identified in the main analyses, i.e., the *HTT* gene with EDS and PSQI for premenopausal women in NEO, and the *ATXN3* gene with sleep duration for the pooled population in NESDA (see Results section). First, linear regression was conducted to assess associations between CAG repeat sizes (in long and short alleles) and baseline IDS score, with the quadratic and interaction terms of alleles to be added if present. Subsequently, for the identified associations of PDAGs and baseline IDS score, we assessed the mediation role of IDS score in the sleep disorders. Mediation analysis based on the causal mediation methodology was performed by using the ‘mediation’ R (version 4.3.1) package (70).

## Acknowledgments

The authors acknowledge all of the participants and investigators in the NEO and NESDA studies. The authors are also thankful to Merel Boogaard for performing the genotyping assays. The NEO study is supported by the participating Departments, Division, and Board of Directors of the Leiden University Medical Center, and by the Leiden University, Research Profile Area Vascular and Regenerative Medicine. The infrastructure for the NESDA study (www.nesda.nl) has been funded through the Geestkracht program of the Netherlands Organisation for Health Research and Development (ZonMw, grant number 10-000-1002) and by participating universities and mental health care organizations (Amsterdam University Medical Centers (location VUmc), GGZ inGeest, Leiden University Medical Center, University Medical Center Groningen, University of Groningen, Lentis, GGZ Friesland, GGZ Drenthe, Dimence, Rob Giel Onderzoekcentrum).

## Funding

L.A. is supported by the China Scholarship Council (CSC; no. 202106240064). H.W. and T.F. were supported by the National Institute of Health (NIH) grant R01HL153814. T.S was supported by NIH grants and R01HL161012 and R01AG080598.

## Author contributions

**L.A.:** Conceptualization, Methodology, Formal analysis, Visualization, Writing-Original Draft, Writing-Review & Editing. **T.F.:** Conceptualization, Methodology, Writing-Original Draft, Writing-Review & Editing, Supervision. **R.N., D.V.H., K.W.V.D. and H.W.:** Conceptualization, Writing-Review & Editing, Supervision. **T.S.:** Methodology, Writing-Review & Editing. **N.A.A., Y.M., B.W.J.H.P., and F.R.R:** Data Curation, Writing-Review & Editing. All authors have reviewed and approved the final version of the manuscript.

## Conflict of interest

The authors declare no competing interest.

## Data availability statement

Due to the privacy of the participants of the included studies and legal reasons, we cannot publicly deposit the data. Data can be made available upon request to interested qualified researchers through the following webpages: NEO: https://www.lumc.nl/en/afdelingen/Clinical-Epidemiology/obesity--related-diseases/ and NESDA: https://www.nesda.nl/researchers/about-nesda/.

## Supplementary Materials

Supplementary methods

Figure S1: Inclusion and exclusion flowchart in NEO

Figure S2: Inclusion and exclusion flowchart in NESDA

Figure S3: Meta-analyses results for sleep duration based on sensitivity analyses

Figure S4: Meta-analyses results for chronotype based on sensitivity analyses

Figure S5. Overall effects of long allele for all observed associations in NEO study

Figure S6. Overall effects of long allele for all observed associations in NESDA study

Table S1: Detailed model results for the identified 31 associations from NEO and NESDA

Table S2: Interaction results for the full conducted interaction test in NEO and NESDA

Table S3: Detailed results of mediation analyses with IDS as the mediator

Supplementary Data 1: Complete regression results for NEO and NESDA

## Notes

### Competing Interest Statement

The authors have declared no competing interest.

### Author Declarations

The The Netherlands Epidemiology of Obesity study was approved by the medical ethical committee of the Leiden University Medical Center. NESDA was approved by the Medical Ethical Committee of the VUmc (reference number 2003/183), and all participants provided written informed consent

