## Supplementary material for "Sex- and Depression-specific Effects of Non-pathogenic CAG Repeats in *HTT*, *ATXN3* and *CACNA1A* on Sleep": Table 1

### Table 1. Characteristics of included study population in NEO and NESDA

| | NEO | | | NESDA at Visit 1 | | | NESDA at Visit 3 | | |
|---|---|---|---|---|---|---|---|---|---|
| | **All** | **Men** | **Women** | **All** | **Men** | **Women** | **All** | **Men** | **Women** |
| N | 3961 | 1935 | 2026 | 2165 | 718 | 1447 | 1788 | 598 | 1190 |
| Age (years), median [IQR] | 56 [51, 61] | 57 [51, 61] | 56 [51, 61] | 44 [31, 54] | 45 [34, 55] | 43 [30, 53] | 45 [32, 55] | 47.5 [36, 57] | 44 [31, 54] |
| Sex = women, n (%) | 2026 (51.1) | - | 2026 (100.0) | 1447 ( 66.8) | 0 (0.0) | 1447 (100.0) | 1190 (66.6) | - | 1190 (100.0) |
| BMI (kg/m$^2$), mean (SD) | 29.70 (4.85) | 29.48 (3.91) | 29.91 (5.60) | 25.45 (4.80) | 26.08 (4.35) | 25.14 (4.98) | 25.62 (4.75) | 26.15 (4.28) | 25.34 (4.96) |
| Menstrual status, n (%) | | | | | | | | | |
| unknown | - | - | - | 18 (0.8) | - | 18 (1.2) | 67 (3.7) | - | 67 (5.6) |
| male respondent | 1935 (48.9) | 1935 (100) | - | 718 (33.2) | 718 (100.0) | 0 (0.0) | 598 (33.4) | 598 (100.0) | 0 (0.0) |
| Premenopause | 775 (19.6) | - | 775 (38.3) | 911 (42.1) | - | 911 (63.0) | 684 (38.3) | - | 684 (57.5) |
| Postmenopause | 1251 (31.6) | - | 1251 (61.7) | 518 (23.9) | - | 518 (35.8) | 439 (24.6) | - | 439 (36.8) |
| IDS score, median [IQR] | 8.0 [5.0, 14.0] | 7.0 [4.0, 11.0] | 10.0 [6.0, 16.0] | 18.0 [9.0, 30.0] | 18.0 [7.0, 30.3] | 19.0 [10.0, 30.0] | 12.0 [6.0, 22.0] | 11.0 [5.0, 22.0] | 12.0 [7.0, 21.0] |
| IDS ≥ 14, n (%) | 1084 (27.4) | 353 (18.2) | 731 (36.1) | 1344 (62.1) | 423 (58.9) | 921 (63.6) | 797 (44.6) | 256 (42.8) | 541 (45.5) |
| Hormone Replacement Therapy = No, n (%) | 1748 (86.3) | - | 1748 (86.3) | - | - | - | - | - | - |
| Sleep Medication = Yes, n (%) | 225 (5.7) | 50 (2.6) | 175 (8.6) | 111 ( 5.1) | 31 ( 4.3) | 80 ( 5.5) | 60 (3.4) | 13 (2.2) | 47 (3.9) |
| **CAG Repeat Sizes** | | | | | | | | | |
| *HTT* long allele (median [IQR]) | 19.0 [18.0, 22.0] | 19.0 [17.0, 22.0] | 19.0 [18.0, 22.0] | 19.0 [17.0, 22.0] | 19.0 [17.0, 21.8] | 19.0 [17.0, 22.0] | 19.0 [17.0, 22.0] | 19.0 [17.0, 22.0] | 19.0 [17.0, 22.0] |
| *HTT* short allele (median [IQR]) | 17.0 [16.0, 17.0] | 17.0 [16.0, 17.0] | 17.0 [16.0, 18.0] | 17.0 [16.0, 17.0] | 17.0 [16.0, 18.0] | 17.0 [16.0, 17.0] | 17.0 [16.0, 17.0] | 17.0 [16.0, 17.0] | 17.0 [16.0, 17.0] |
| *CACNA1A* long allele (median [IQR]) | 13.0 [12.0, 13.0] | 13.0 [12.0, 13.0] | 13.0 [12.0, 13.0] | 13.0 [12.0, 13.0] | 13.0 [12.0, 13.0] | 13.0 [12.0, 13.0] | 13.0 [12.0, 13.0] | 13.0 [12.0, 13.00] | 13.0 [12.0, 13.0] |
| *CACNA1A* short allele (median [IQR]) | 11.0 [11.0, 12.0] | 11.0 [11.0, 12.0] | 11.0 [11.0, 12.0] | 11.0 [11.0, 12.0] | 11.0 [11.0, 12.0] | 11.0 [11.0, 12.0] | 11.0 [11.0, 12.0] | 11.0 [11.0, 12.0] | 11.0 [11.0, 12.0] |
| *ATXN3* long allele (median [IQR]) | 23.0 [23.0, 27.0] | 23.0 [23.0, 27.0] | 23.0 [23.0, 27.0] | 23.0 [23.0, 27.0] | 23.0 [23.0, 27.0] | 23.0 [23.0, 27.0] | 23.0 [23.0, 27.0] | 23.0 [23.0, 27.0] | 23.0 [23.0, 27.0] |
| *ATXN3* short allele (median [IQR]) | 21.0 [14.0, 23.0] | 20.0 [14.0, 23.0] | 21.0 [14.0, 23.0] | 20.0 [14.0, 23.0] | 20.0 [14.0, 23.0] | 20.0 [14.0, 23.0] | 20.0 [14.0, 23.0] | 20.0 [14.0, 23.0] | 20.0 [14.0, 23.0] |
| **Sleep Traits** | | | | | | | | | |
| PSQI, median [IQR] | 4.0 [3.0, 6.0] | 4.0 [2.0, 5.0] | 5.0 [3.0, 8.0] | | | | | | |

| | | | | | | | | | |
|---|---|---|---|---|---|---|---|---|---|
| EDS = YES, n (%) | 999 (25.2) | 495 (25.6) | 504 (24.9) | | | | | | |
| Sleep duration (hour), mean (SD)* | 7.98 (1.09) | 7.68 (1.08) | 8.26 (1.02) | | | | 7.75 (0.98) | 7.46 (0.88) | 7.90 (1.00) |
| WHIIRS, mean (SD) | | | | 7.99 (5.07) | 7.69 (5.13) | 8.13 (5.04) | | | |
| WHIIRS $\geq$ 9, Yes, n (%) | | | | 913 ( 42.2) | 292 ( 40.7) | 621 ( 42.9) | | | |
| Chronotype, n(%) | | | | | | | | | |
| Morning type | 876 (22.1) | 491 (25.4) | 385 (19.0) | | | | 354 (19.8) | 127 (21.2) | 227 (19.1) |
| Neither type | 2314 (58.4) | 1051 (54.3) | 1263 (62.3) | | | | 868 (48.5) | 278 (46.5) | 590 (49.6) |
| Evening type | 771 (19.5) | 393 (20.3) | 378 (18.7) | | | | 566 (31.7) | 193 (32.3) | 373 (31.3) |

*: In NESDA, the outcome of sleep duration is available in N=1333 (Men N = 455, Women N = 878).

Abbreviations: BMI, body mass index; CAG, cytosine–adenine–guanine; EDS, Epworth Daytime Sleepiness; IQR, interquartile range; IDS, Inventory for Depressive Symptomatology; PSQI, Pittsburgh Sleep Quality Index; SD, standard deviation; WHIIRS, Women's Health Initiative Insomnia Rating Scale.

**Table 2. Significant associations between PDAG CAG repeat sizes and the dichotomized Inventory for Depressive Symptomatology score**

| Study | PDAGs | Allele | Estimate | Standard error | *P* value |
|---|---|---|---|---|---|
| NEO | | | | | |
| | *HTT* | Long allele | -0.050 | 0.037 | 0.179 |
| | *HTT* | Short allele | 0.137 | 0.056 | 0.015 |
| NESDA | | | | | |
| | *ATXN3* | Long allele | 0.046 | 0.017 | 0.008 |
| | *ATXN3* | Short allele | -0.008 | 0.014 | 0.581 |

NEO analysis was performed in the premenopausal subgroup. NESDA analysis included the pooled population.
